# Improving estimation of Parkinson’s disease risk - The enhanced PREDICT-PD algorithm

**DOI:** 10.1101/2020.03.16.20037002

**Authors:** Jonathan P. Bestwick, Stephen D. Auger, Cristina Simonet, Richard N. Rees, Daniel Rack, Mark Jitlal, Gavin Giovannoni, Andrew J. Lees, Jack Cuzick, Anette E. Schrag, Alastair J. Noyce

**Author notes:** These authors contributed equally to this work (Jonathan P. Bestwick, Stephen D. Auger, Anette E. Schrag, Alastair J. Noyce). Corresponding author: Dr Alastair Noyce, Preventive Neurology Unit, Wolfson Institute of Preventive Medicine, Queen Mary University of London, Charterhouse Square, London, EC1M 6BQ. Funding sources for study: Parkinson’s UK (G1606).

## Abstract

**Background:** We previously reported a basic algorithm to identify risk of Parkinson’s disease (PD) using published data on risk factors and prodromal features. Using this algorithm, the PREDICT-PD study identified individuals at increased risk of PD, using tapping speed, hyposmia and REM sleep behaviour disorder (RBD) as “intermediate” markers of prodromal PD until sufficient incident cases accrued.

**Objectives:** To develop and test an enhanced algorithm which incorporates these intermediate markers into the risk model.

**Methods:** Intermediate markers were incorporated into risk estimates, and likelihood ratios for both the presence and absence of risk factors were used. Risk estimates were compared using the enhanced and the basic algorithm in members of the PREDICT-PD pilot cohort.

**Results:** The enhanced PREDICT-PD algorithm yielded a much greater range of risk estimates than the basic algorithm (346 - 784-fold difference between the 10^th^ and 90^th^ centiles vs 10 - 12-fold respectively). There was a greater increase in risk of PD with increasing risk scores for the enhanced algorithm than for the basic algorithm (hazard ratios per one standard deviation increase in log risk of 2.73 [95% CI 1.68 – 4.45; p<0.001] versus 1.47 [95% CI 0.86 - 2.51; p=0.16] respectively). Estimates from the enhanced algorithm also correlated more closely with subclinical striatal DaT-SPECT dopamine depletion (R^2^=0.14, p=0.010 vs R^2^=0.043, p=0.17).

**Conclusions:** Incorporating the previous intermediate markers of prodromal PD and using likelihood ratios improved the accuracy of the PREDICT-PD prediction algorithm for PD.

## INTRODUCTION

Neurodegeneration preceding a formal diagnosis of Parkinson’s disease (PD) is associated with identifiable motor and non-motor features. Evidence-based algorithms have been developed to try to identify individuals in this pre-diagnostic phase according to exposure to common risk factors and with simple screening tests. Two notable approaches to risk estimation are the PREDICT-PD algorithm^1^ and the MDS prodromal PD research criteria.^2^

The PREDICT-PD study is a prospective study in 60 to 80 year olds (the “PREDICT-PD pilot cohort”)^3^, specifically designed to estimate risk from information gathered using simple online tests and remotely administered screening tools; this includes demographic information, environmental exposures and early symptoms identified in a systematic review and meta-analysis.^4^ Reduced finger tapping speed on a keyboard tapping task, hyposmia and probable REM sleep behaviour disorder (RBD) were used as “intermediate” markers for prodromal PD. The MDS criteria, published in 2015, incorporate additional clinical and radiological tests, including these three factors.

As participants of the ongoing longitudinal PREDICT-PD pilot cohort develop PD, it is now possible to use incident diagnosis of PD as an outcome, and improve the algorithm by incorporating intermediate markers into the risk estimates. In addition, PREDICT-PD risk estimation has previously been based upon odds ratios. This had the limitation that if a risk factor is known to be absent, there is no adjustment to the change in risk this represents. Using likelihood ratios instead allows better characterisation of overall risk, as the presence (LR+) or absence (LR-) of each individual marker is considered in the algorithm.

Here, we sought to refine the PREDICT-PD algorithm, first by changing the methods of risk estimation from odds ratios to likelihood ratios. We then also incorporated the assessment of tapping speed, smell and probable RBD into risk estimates. To assess whether this provided improved risk estimation, we compared the distributions of risk derived from the enhanced algorithm to those using the basic PREDICT-PD algorithm, and also to the MDS criteria algorithm.^2,5^ We then considered the members of the PREDICT-PD pilot cohort who have developed PD to date and assessed the algorithm’s risk estimation preceding formal diagnosis. We also assessed the relationship between risk estimates and subclinical striatal dopamine depletion measured by dopamine transporter imaging (DaT-SPECT) in a sub-group of the participants.

## METHODS

### Creating an enhanced PREDICT-PD algorithm

Previous PREDICT-PD risk estimation has been based upon single odds ratios, with risk estimates only adjusted if a risk factor was known to be present. To select the most appropriate likelihood ratios to include in an “enhanced” version of the PREDICT-PD algorithm, we first sought the best available evidence in the published literature. Berg and colleagues^2^ have previously published likelihood ratios for a number of PD risk factors which were included in this calculation. However, a number of risk factors included in PREDICT-PD risk estimation had no previous reported positive and negative likelihood ratios for their association with PD. Namely: head injury and the use of non-steroidal anti-inflammatory medications (NSAIDs), calcium channel blockers (CCB), beta blockers and alcohol. For these, we converted the previously used odds ratios into likelihood ratios using prevalence data from the East London Primary Care database held by the Clinical Effectiveness Group at Queen Mary, University of London (n = 1,016,277) with the following formulae:

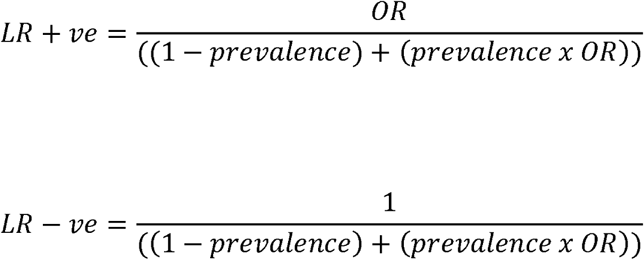

After noting that the equation specifying the age-specific risk in the PREDICT-PD algorithm was underestimating risk at older ages (based on a regression of risk against age^1^), we examined whether using the risks based on categories of age in the MDS-criteria (which provided more data points to base the regression equation on than previously used) would lead to a better fit.

We further sought to enhance the PREDICT-PD algorithm’s risk estimation by including motor (alternate finger tapping) and non-motor (objective assessment of smell and subjective RBD) intermediate markers in the risk score. Motor function was determined using the BRAIN tap test.^6^ Smell was assessed using the University of Pennsylvania Smell Identification Test (UPSIT).^7^ RBD was assessed using the REM-sleep behaviour disorder questionnaire (RBDSQ).^8^ In parallel work, likelihood ratios for smell were calculated according to logistic regression models of: (i) the 16 odours identified that were shown to be associated with PD and (ii) a subset of the 6 odours that were most strongly associated with PD; from the bivariate Gaussian distributions of delta kinesia score (KS) and akinesia time (AT) multiples of median (MoM) values in PD patients and controls; and from the 6 questions of the RBDSQ that were shown to be associated with PD.^9^ The updated PREDICT-PD risk estimates (incorporating these three factors and using likelihood ratios rather than odds ratios) are referred to as the “enhanced” PREDICT-PD algorithm henceforth, whereas scores without inclusion of the intermediate markers and based upon odds ratios are referred to as the “basic” PREDICT-PD algorithm.

### Data collection

The PREDICT-PD pilot cohort comprised 1,323 healthy 60 to 80 -year olds at baseline (mean age 67.2 years, SEM 0.13, 60.9% females). Participants have completed annual online surveys and a keyboard-tapping task up to 6 times over 7 years. For the 5th year of follow-up, no data were collected due to a change in the study website. Detailed information about the recruitment process and data collection methods are described elsewhere^1,3^. If there were missing data in one year but available data in subsequent years, we imputed the missing years’ data by assuming risk exposures to be the same as those in the preceding year’s follow-up, but accounting for the fact that they would be one year older (which is associated with a small increase in risk). Similarly smell testing was only done at baseline and in year 3, so the results of smell tests at baseline were applied to risk calculations in years 1 and 2, and results of smell tests in year 3 were applied to risk calculations in years 4 and 6. For those for whom no further follow-up data were available, we did not impute any missing data as we could not be certain of continued consent.

### Comparison of risk scores

For each survey year, histograms of risk estimate for each algorithm (basic PREDICT-PD, enhanced PREDICT-PD and MDS criteria) were generated and selected centiles of risk (1^st^, 2.5^th^, 10^th^, 25^th^, 50^th^, 75^th^, 90^th^, 95^th^, 97.5^th^ and 99^th^) were calculated. Risks were expressed as odds and presented graphically on a log scale. We examined the fold difference in risk between the 10^th^ and 90^th^ centile, and the 25^th^ and 75^th^ centile. We did not consider the fold difference in risk between the minimum and maximum, or for example, the 1^st^ and 99^th^ centile as this would be subject to bias from outlying risk estimates or give unstable estimates of fold differences, which is particularly relevant given the inclusion of continuous variables in risk scores (age and the delta KS/MoM AT parameters). Cox proportional hazard models were used to determine the association between baseline risk scores using each algorithm and incident PD.

### Comparison of risk estimates with DaT-SPECT binding

46 people in the PREDICT-PD pilot cohort had DaT-SPECT imaging. The methods relating to how these images were acquired have been described elsewhere.^11,12^ None of the 46 individuals had been diagnosed with PD during continued follow-up. We sought to investigate whether risk estimates were be related to striatal dopamine binding. Linear regression was used to investigate the relationship between risk estimates (expressed as log odds) according to each algorithm with each individual’s corresponding striatal DaT-SPECT binding data. Each participant had bilateral DAT-SPECT binding values and we took the lower of the two values as a marker of potential dopamine depletion. The risk estimates closest in time to the DaT-SPECT imaging were used in the analysis, which were either year 2 or year 3 risks.

### Ethical approval

The PREDICT-PD study was approved by Central London Research Committee 3 (reference number 10/H0716/85)

## RESULTS

### The enhanced PREDICT-PD algorithm

The new likelihood ratios for risk factors which had no previously reported positive and negative likelihood ratios for their association with PD are presented in Table 1, together with the prevalence and odds ratio data used to make these calculations. All the factors which were available for inclusion into risk estimates in either the basic or enhanced PREDICT-PD risk estimates are outlined in Table 2, alongside the most recent MDS research criteria likelihood ratios. No prevalence data were available for pesticide exposure or having a 1^st^ degree relative with PD, so negative likelihood ratios for these factors could not be calculated.

**Table 1:** Prevalence, odds ratio and likelihood ratios for a positive (LR+) or negative (LR-) association with PD for risk factors without previous estimates

| Risk factor | Prevalence | Odds Ratio | LR+ | LR- |
| --- | --- | --- | --- | --- |
| Head injury | 0.03 | 1.58 | 1.55 | 0.98 |
| NSAID use | 0.8 | 0.83 | 0.96 | 1.16 |
| CCB use | 0.428 | 0.9 | 0.94 | 1.04 |
| Beta blocker use | 0.284 | 1.28 | 1.19 | 0.93 |
| Alcohol use | 0.8 | 0.9 | 0.98 | 1.09 |

**Table 2:** Likelihood ratios (LRs) and Odds Ratios (ORs) for PD risk factors collected in the pilot PREDICT-PD cohort using the PREDICT-PD risk estimation algorithms and MDS research criteria

| Factor | MDS (LRs) | Basic PREDICT (ORs) | Enhanced PREDICT (LRs) |
| --- | --- | --- | --- |
| Factors in both MDS and PREDICT algorithms |  |  |  |
| Age | Categorical 5-year age intervals | Age-based equation | Age-based equation |
| Sex | male 1.2, female 0.8 | female 0.67 | male 1.2, female 0.8 |
| Coffee use | LR+ = 0.88, LR- = 1.35 | 0.67 | LR+ = 0.88, LR- = 1.35 |
| Current smoker | LR+ = 0.51 | 0.44 | LR+ = 0.51 |
| Former smoker | LR+ = 0.91 | 0.78 | LR+ = 0.91 |
| 1st degree relative | LR+ = 2.5 | 3.2 | LR+ = 2.5 |
| Constipation | LR+ = 2.5, LR- = 0.82 | 2.3 | LR+ = 2.5, LR- = 0.82 |
| Erectile Dysfunction | LR+ = 3.4, LR- = 0.87 | 3.8 | LR+ = 3.4, LR- = 0.87 |
| Depression/anxiety | LR+ = 1.6, LR- = 0.88 | 1.86 | LR+ = 1.6, LR- = 0.88 |
| Factors in one algorithm but not the other |  |  |  |
| Objective motor impairment* | LR+ = 3.5, LR- = 0.60 | - | Bivariate Gaussian model based equation** |
| REM-sleep behaviour disorder* | LR+ = 2.8, LR- = 0.89 | - | Logistic regression model based equation** |
| Olfactory impairment* | LR+ = 4.0, LR- = 0.43 | - | Logistic regression model based equation** |
| Pesticides exposure | LR+ = 1.5 | - | LR+ = 1.5 |
| Never smoked | LR+ = 1.2 | - | LR+ = 1.25 |
| Head injury | - | 1.58 | LR+ = 1.55, LR- = 0.098 |
| NSAID use | - | 0.83 | LR+ = 0.96, LR- = 1.16 |
| CCB use | - | 0.9 | LR+ = 0.94, LR- = 1.04 |
| Beta blocker use | - | 1.28 | LR+ = 1.19, LR- = 0.93 |
| Alcohol | - | 0.9 | LR+ = 0.98, LR- = 1.09 |

\*Previously used as intermediate outcome markers and so did not feature in the basic PREDICT-PD algorithm.
\*\*Bestwick et al. 2020<sup>9</sup>

Supplementary Figure 1 shows the risk (expressed as an odds) according to age in the basic PREDICT-PD algorithm and the revised risk according to age in the enhanced PREDICT-PD algorithm (based on the risk according to age categories in the MDS criteria).^2,5^ The revised equation for estimating age-related risk is:

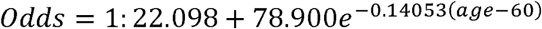

Between the ages of 60 and 70, the old and revised equations estimate near identical risk. There is some difference at higher ages however, for example the revised equation gives a risk for an 80-year old of 1:27, whereas the previous equation gave a risk of 1:31.

### Distributions of risk

Figure 1 shows histograms of risk (expressed as odds) at baseline and year 6. Supplementary Figure 2 shows histograms for all years. Shown in the figures are the distributions using the basic PREDICT-PD algorithm, the enhanced PREDICT-PD algorithm (using either the most discriminant 16 or 6-items from the 40-item UPSIT) and the MDS criteria algorithm. Supplementary Table 1 shows selected centiles of risk for each algorithm and survey year. Figure 1 (and Supplementary Figure 2) shows that for each survey year, the basic PREDICT-PD algorithm yields the least spread in risk, with the enhanced PREDICT-PD algorithm producing greater spread (with similar spread whether the 16-item or the 6-item smell test subset were used for risk estimation). Over 6 time points, there was between a 10 and 12-fold difference in risk from the 10^th^ to 90^th^ centile of risk for the basic PREDICT-PD algorithm, compared to between a 346 and 784-fold difference in risk for the enhanced PREDICT-PD algorithm using a 16-item smell test, between a 217 and 522-fold difference in risk for the enhanced PREDICT-PD algorithm using a 6-item smell test, and between a 35 and 48-fold difference in risk for the MDS criteria algorithm. Between the 25^th^ and 75^th^ centile of risk, the respective fold differences were between 3.5 and 4.3, between 26 and 40, between 22 and 28, and between 6.7 and 9.1. Supplementary Table 1 provides risks for a greater range of centiles.

**Figure 1:**
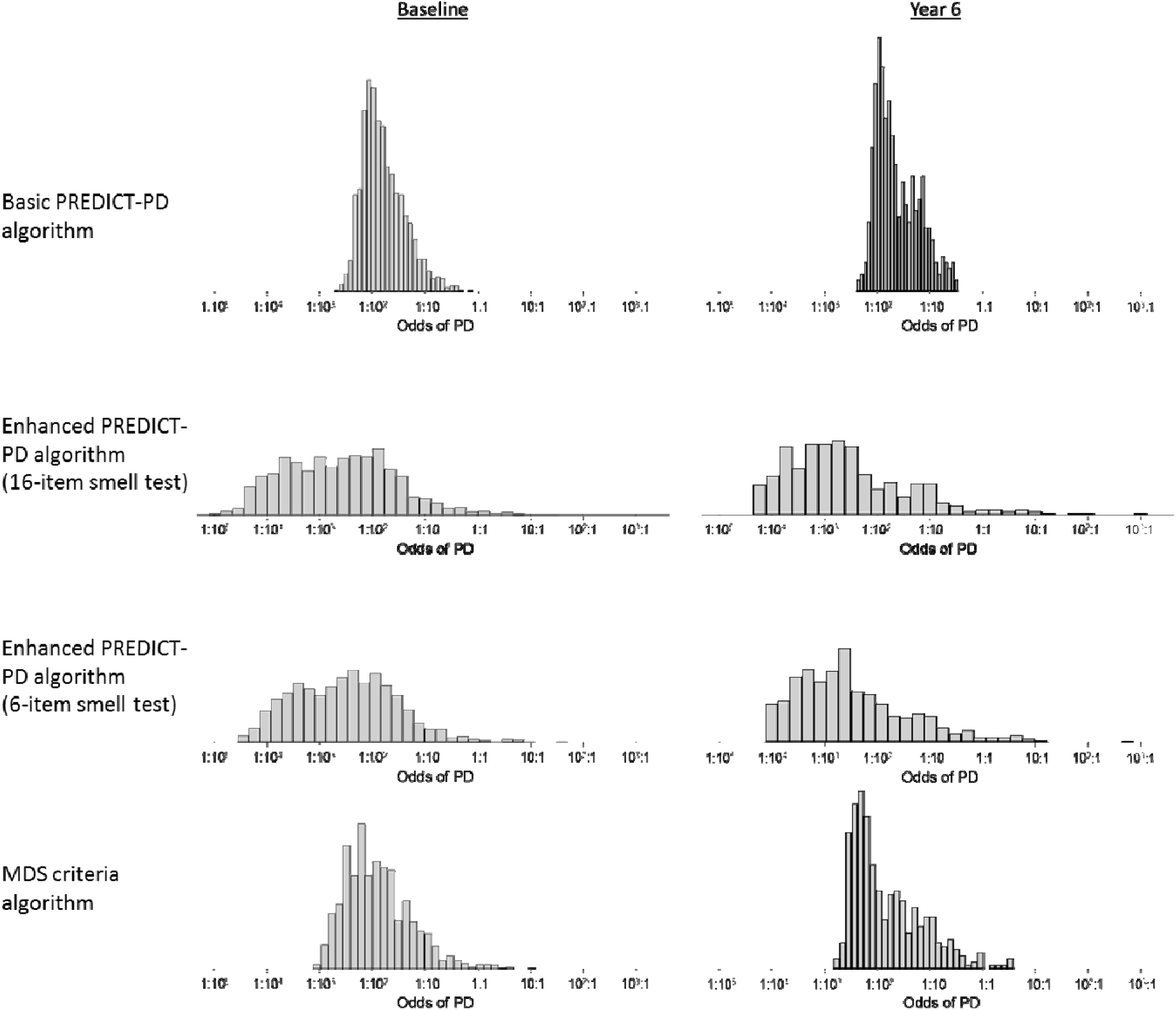
Histograms of risk scores for PREDICT-PD participants (presented as odds) at baseline (far left) and in the latest survey year with the basic PREDICT-PD algorithm (first row), the enhanced PREDICT-PD algorithm using a 16 item smell test (second row), the enhanced PREDICT-PD algorithm using a 6-item smell test (third row) and the MDS criteria algorithm (third row).

### Comparison of PREDICT-PD risk scores in participants diagnosed with PD

Ten people in the PREDICT-PD cohort have been diagnosed with PD to date. Table 3 shows the relationship between baseline risk score and incident PD using a Cox proportional hazards model. For both a 10-fold increase in risk and a one standard deviation increase in log risk, the hazard ratios for each risk algorithm were similar; but whilst both the enhanced PREDICT-PD (using the 16-item smell test or the 6-item smell test) and the MDS criteria algorithm had statistically significant associations with an increased risk of PD (p<0.001, p=0.002 and p=0.001 respectively), the association for the basic PREDICT-PD score did not reach significance (p=0.157). The enhanced PREDICT-PD algorithms and MDS criteria algorithm gave similar results, but it should be noted that only four of the ten incident PD cases had UPSIT scores at baseline, so for the remaining six, UPSIT scores were not included in the risk estimates. Furthermore, one of the six incident PD cases who did not have an UPSIT score at baseline also did not complete the BRAIN test at baseline and could not have objective motor impairment included in calculation of risk estimates.

**Table 3:** Hazard ratios (HR) of incident PD at 6 years of follow-up for a 10-fold increase in baseline risk and a one standard deviation (SD) increase in baseline log risk according to risk algorithm.

| Algorithm | HR per 10-fold increase<br>in risk (95% CI) | HR per SD of log risk<br>(95% CI) | p-value |
| --- | --- | --- | --- |
| Basic PREDICT-PD | 2.58 (0.69-9.56) | 1.47 (0.86-2.51) | 0.157 |
| Enhanced PREDICT-PD (16-item smell test) | 2.75 (1.69-4.47) | 2.91 (1.74-4.85) | <0.001 |
| Enhanced PREDICT-PD (6-item smell test) | 2.81 (1.70-4.63) | 2.73 (1.68-4.45) | <0.001 |
| MDS criteria | 3.12 (1.55-6.31) | 2.05 (1.32-3.19) | 0.001 |

### Comparison of risk estimates with DaT-SPECT binding

Figure 2 shows the relationship between risk using each algorithm and striatal dopamine binding ratios with DaT-SPECT imaging in a subgroup of 46 individuals in the PREDICT-PD cohort, none of whom have been diagnosed with PD. The figure shows that for each algorithm striatal binding values were lower as risks became higher; this relationship was not statistically significant for the basic PREDICT-PD algorithm (R^2^=0.043, p=0.165) but was for the enhanced PREDICT-PD algorithm using the 16-item smell test (R^2^=0.141, p=0.010), the enhanced PREDICT-PD algorithm using the 6-item subset (R^2^=0.103, p=0.030) and the MDS criteria algorithm (R^2^=0.165, p=0.005) indicating that the enhanced PREDICT-PD and MDS criteria algorithm’s risk estimates bear closer relation to striatal dopaminergic depletion, even in individuals who do not have a clinical PD diagnosis.

**Figure 2:**
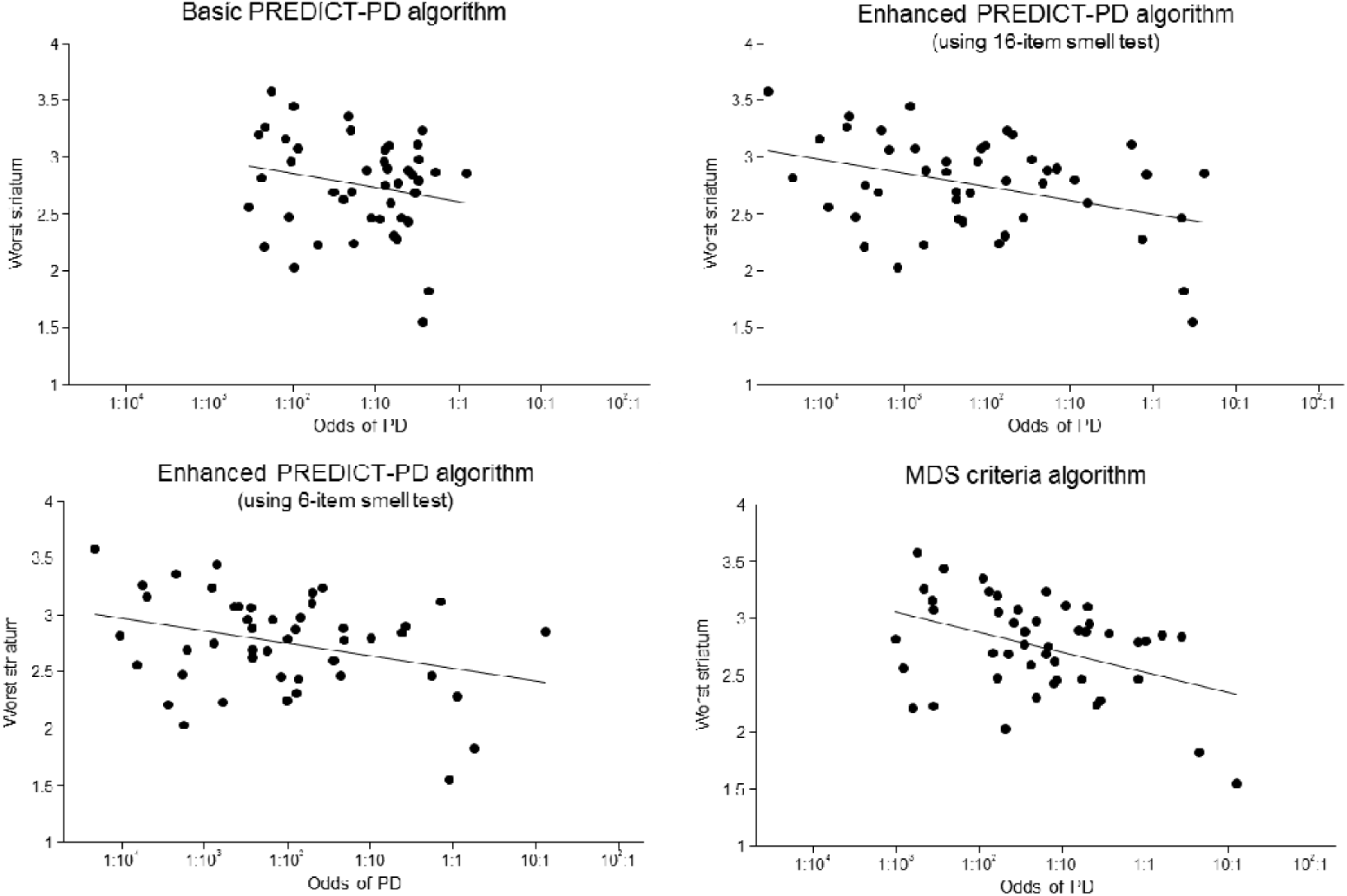
Regression analysis comparing risk estimates (expressed as odds on a log scale) according to algorithm against levels of striatal dopamine depletion according DaT-SPECT imaging data.

## DISCUSSION

Despite just 10 people in the PREDICT-PD pilot cohort receiving a formal diagnosis of PD, the difference in relationship between baseline risk and incident PD between the basic and enhanced PREDICT-PD algorithms, and the much greater range of risk attributable to the enhanced PREDICT-PD algorithm suggests a meaningful improvement in risk estimation. Further evidence of the enhanced algorithm’s ability to identify those at risk of subsequent PD comes from the analysis of DaT-SPECT data, where the enhanced algorithm’s risk estimates bore a significant and closer relation to striatal dopamine depletion even when clinical PD is not evident. As the cohort expands to a target of 10,000 participants, more data accrues for incident cases and at a longer time prior to a diagnosis of PD, the true temporal performance of the enhanced risk prediction will become clearer.

The main alternative to the PREDICT-PD approach to risk estimation is that of the MDS research criteria^2^. There is a suggestion that the enhanced PREDICT-PD algorithm has the potential to perform better than the MDS criteria algorithm since a greater range of risks estimates are likely to lead to greater discrimination between those who do and do not develop PD, although the number of PD cases in the PREDICT-PD pilot cohort is so far too small to confirm this. The main differences between how the two algorithms derive risk estimates are in the range of risk factors included and how risk according to age, motor impairment, RBD and smell are defined. The MDS criteria determines age-related risk according to which 5-year age interval a person falls into whereas the enhanced PREDICT-PD algorithm uses a continuous function to assign risk according to exact age, because categorising a continuous variable leads to a loss of information.^11^ Similarly, for smell, RBD and motor impairment, the MDS criteria use the binary presence or absence of a risk factor, rather than identifying specific items in the UPSIT or RBDSQ that are predictive of PD and generating likelihood ratios accordingly, or by treating motor impairment as a continuous variable as were done in the enhanced PREDICT-PD algorithm. The MDS criteria do include a wider range of radiological and clinical measures which provide important information but require more intensive monitoring than the simple, remotely administered tests used in PREDICT-PD. The MDS criteria do not however, consider head injury or the use of NSAIDs, calcium channel blockers, beta blockers or alcohol. We propose these are valuable additions. A history of head injury might be particularly relevant: systematic review found the odds ratio for head injury in PD to be 1.58.^11^ Another difference between the MDS and PREDICT-PD approaches is their respective reporting of total combined risk in terms of percentage probability and odds (not to be confused with odds *ratio* of individual factors). It is possible to convert between the two, but here we favour expressing risk in terms of odds due to the resultant lower skew when plotting distributions on a log scale. Odds also has the advantage of allowing larger fold-changes between the highest and lowest risks to be demonstrated; their range is infinite rather than the bounding of percentage probabilities between 0 and 100.

In the enhanced PREDICT-PD algorithm, we used the 16 smells that are present in both the US and UK UPSIT are predictive of PD case status, and a subset of the 6 odours most strongly associated with PD.^9^ While the enhanced PREDICT-PD algorithm with the 16 items led to the greatest range of risks, the enhanced PREDICT-PD algorithm with the 6 item test led to greater range of risks than the basic PREDICT-PD algorithm and the MDS criteria algorithm and the cost savings may outweigh the increased range of risks versus either the full 40-item UPSIT or a 16 item test.

A limitation of both approaches is that, to date, these probability-based algorithms have largely been derived from retrospective studies and systematic review of risk factors in people already diagnosed with PD. As a result, the PREDICT-PD and MDS estimation models currently assume a level of independence of risk factors which is unlikely to be true in reality. Models such as PREDICT-PD or the MDS criteria act as a first approximation to risk estimation but will need to be refined once prospective observational cohorts, such as the PREDICT-PD, the PARS^12^ and the Bruneck^13^ studies mature and provide more information regarding correlations between risk factors in the years preceding formal diagnosis with PD. This might allow risk estimation algorithms to evolve from taking a univariate approach, where individual risk factors are considered in isolation, to more sophisticated analysis of multivariate patterns. At present, there are insufficient high-quality data to allow for this to be done effectively. Further improvement could also come from considering individual risk factors in greater detail. A greater number of risk factors could be considered in terms of more than just a binary presence or absence of risk. More information such as the age at onset of PD in a first degree relative, pack years of smoking or severity of constipation and depression would be likely to provide useful information with regards PD risk. New risk associations are also continually being discovered for as diverse a range of factors as dietary preferences, type 2 diabetes, personality traits or history of migraine or epilepsy.^14^ Algorithms estimating risk will need to continually update in the presence of relevant new information, as has recently been the case with the MDS criteria.^5^

## Conclusion

Probability-based algorithms offer an effective means of identifying people at highest risk of developing PD with simple, remotely administered tests. This allows for the recruitment of very large sample sizes, while identifying individuals who can be targeted for closer monitoring and investigation with more resource-intensive tests. This could be particularly valuable in a research setting, where the low incidence of PD complicates prospective investigation in the years prior to development of overt clinically diagnosable PD. The participants providing data in the right tail of the histograms in Figure 1 are of particular interest for targeting more intensive testing and follow-up. It may be that these individuals estimated to be at highest risk could allow identification of pre-diagnostic biomarkers for PD. This is supported by the evidence from the DaT-SPECT data that higher risk estimates are associated with greater striatal dopamine depletion.

As large prospective cohorts mature, a greater understanding of relationships between pre-diagnostic features with more intensive investigation in high-risk individuals could also provide important information for further refining risk estimation.

## Data Availability

It is the intention of the authors to make study data
available for sharing

## ACKNOWLEDGMENTS

We would like to thank all the participants in the PREDICT-PD pilot cohort and Zoheb Shah.

## AUTHORS’ ROLES

1) Research project: A. Conception, B. Organization, C. Execution; 2) Statistical Analysis: A. Design, B. Execution, C. Review and Critique; 3) Manuscript: A. Writing of the first draft, B. Review and Critique.

J.P.B.: 1A, 1B, 2A, 2B, 2C, 3B

S.D.A.: 2A, 2B, 2C, 3A, 3B

C.S.: 3B

R.R.: 3B

D.R.: 2B, 3B

M.J.: 2A, 2B, 2C, 3B

G.G.: 1A, 1B, 3B

A.J.L.: 1A, 1B, 3B

J.C.: 2C, 3B

A.S.: 1A, 1B, 1C, 2A, 2C, 3B

A.J.N.: 1A, 1B, 1C, 2A, 2B, 2C, 3B

## REFERENCES

1. Noyce AJ, Bestwick JP, Silveira-Moriyama L, et al. PREDICT-PD: identifying risk of Parkinson’s disease in the community: methods and baseline results. J Neurol Neurosurg Psychiatry. 2014;85(1):31–37. doi:10.1136/JNNP-2013-305420

2. Berg D, Postuma RB, Adler CH, et al. MDS research criteria for prodromal Parkinson’s disease. Mov Disord. 2015;30(12):1600–1611. doi:10.1002/mds.26431

3. Noyce AJ, R’Bibo L, Peress L, et al. PREDICT-PD: An online approach to prospectively identify risk indicators of Parkinson’s disease. Mov Disord. 2017;32(2):219–226. doi:10.1002/mds.26898

4. Noyce AJ, Bestwick JP, Silveira-Moriyama L, et al. Meta-analysis of early nonmotor features and risk factors for Parkinson disease. Ann Neurol. 2012;72(6):893–901. doi:10.1002/ana.23687

5. Heinzel S, Berg D, Gasser T, Chen H, Yao C, Postuma RB. Update of the MDS research criteria for prodromal Parkinson’s disease. Mov Disord. August 2019:mds.27802. doi:10.1002/mds.27802

6. Noyce AJ, Nagy A, Acharya S, et al. Bradykinesia-akinesia incoordination test: validating an online keyboard test of upper limb function. PLoS One. 2014;9(4):e96260. doi:10.1371/journal.pone.0096260

7. Doty RL, Shaman P, Dann M. Development of the University of Pennsylvania Smell Identification Test: a standardized microencapsulated test of olfactory function. Physiol Behav. 1984;32:489–502

8. Stiasny-Kolster K, Mayer G, Schäfer S, Möller JC, Heinzel-Gutenbrunner M, Oertel WH. The REM sleep behavior disorder screening questionnaire-A new diagnostic instrument. Mov Disord. 2007;22(16):2386–2393. doi:10.1002/mds.21740

9. Bestwick JP, Auger SD, Schrag AE et al. Maximising information on smell, quantitative motor impairment and probable REM-sleep behaviour disorder in the prediction of Parkinson’s disease. medRxiv 2020; doi: https://doi.org/10.1101/2020.03.03.20023994

10. Noyce AJ, Dickson J, Rees RN, et al. Dopamine reuptake transporter–single-photon emission computed tomography and transcranial sonography as imaging markers of prediagnostic Parkinson’s disease. Mov Disord. 2018. doi:10.1002/mds.27282

11. Soderlund TA, Dickson JC, Prvulovich E, et al. Value of Semiquantitative Analysis for Clinical Reporting of 123I-2- -Carbomethoxy-3 -(4-Iodophenyl)-N-(3-Fluoropropyl)Nortropane SPECT Studies. J Nucl Med. 2013. doi:10.2967/jnumed.112.110106

12. Siderowf A, Jennings D, Eberly S, et al. Impaired olfaction and other prodromal features in the Parkinson At-Risk Syndrome study. Mov Disord. 2012;27(3):406–412. doi:10.1002/mds.24892

13. Mahlknecht P, Gasperi A, Djamshidian A, et al. Performance of the Movement Disorders Society criteria for prodromal Parkinson’s disease: A population-based 10-year study. Mov Disord. 2018;33(3):405–413. doi:10.1002/mds.27281

14. Heilbron K, Noyce A, Fontanillas P, et al. The Parkinson’s Phenome: Traits Associated with Parkinson’s Disease in a Large and Deeply Phenotyped Cohort. bioRxiv. February 2018:270934. doi:10.1101/270934

